# Mental health of undocumented college students during the COVID-19 pandemic

**DOI:** 10.1101/2020.09.28.20203489

**Authors:** Jarid Goodman, Sharron Xuanren Wang, Rubi A. Guadarrama Ornelas, Marina Hernandez Santana

## Abstract

The coronavirus disease 2019 (COVID-19) pandemic has caused a surge in mental health problems across the United States, and some reports suggest a more severe impact for racial and ethnic minorities. The present study was conducted to gain a preliminary understanding of the mental health consequences of the COVID-19 pandemic specifically for dreamers, i.e., undocumented immigrants who entered the U.S. as minors. A population of about 150 dreamers currently enrolled at a public university in Delaware were invited to participate in an online survey. The survey contained questions about demographics, mental health, academics, immigration, COVID-19 infection, and unemployment, in addition to mental health screens for anxiety (GAD-7), depression (PHQ-9), and stress (PSS-10). In total, 109 dreamers completed the survey. We observed remarkably high clinical levels of anxiety and depression: 47% of the dreamers met the clinical cutoff for anxiety, 63% met the cutoff for depression, and 67% (2 in 3) met the cutoff for anxiety and/or depression. Rates of anxiety and depression in our sample were significantly higher than those recently reported for college students overall, suggesting that dreamers may be experiencing a more severe mental health impact of the COVID-19 pandemic. We also found that pandemic-induced concerns about finances, COVID-19 infection, immigration, and unemployment (among other factors) were associated with greater anxiety, stress, and depression among the dreamers in our sample. The present findings are consistent with recent predictions by social scientists that the COVID-19 pandemic would have a disproportionately negative impact on the mental health of undocumented immigrants.

**Highlights:**

- Nearly half the dreamers (47%) met the clinical cutoff for anxiety, and 62% met the clinical cutoff for depression.
- 2 in 3 dreamers met the clinical cutoff for anxiety and/or depression.
- The percentage of dreamers meeting the cutoff for anxiety (47%) and depression (63%) were significantly higher than observed for college students overall during the pandemic (31% and 41%, respectively).
- The percentage of dreamers meeting the cutoff for anxiety was also significantly higher than previously observed for undocumented college students in a 2015 survey (35%).
- 60% of dreamers said the pandemic had a serious negative impact on their mental health, while 90% said the pandemic made them more anxious about finances.
- 90% of dreamers said the pandemic made it harder for them to concentrate on coursework, and 2 in 3 said pandemic-related anxiety hurt their academic performance.
- About 1 in 3 dreamers are “extremely worried” that the pandemic will prevent them from achieving their academic and professional goals.
- 76% of dreamers said the pandemic increased their fears of DACA termination.
- 10% of dreamers said they or an immediate family member suspected COVID-19 infection at some point but did not get tested for fear of detainment or deportation.
- About 1 in 5 dreamers said they would be “extremely worried” to seek treatment or have a family member seek treatment for COVID-19 due to fears of detainment or deportation.
- Dreamers who reported one or both parents lost their job due to the pandemic had significantly greater anxiety and depression scores and were more likely to meet clinical cutoffs for anxiety and depression.

## Introduction

The coronavirus disease 2019 (COVID-19) pandemic has had a dramatic negative impact on several facets of our collective wellbeing, including mental health. There is extensive evidence of increased mental health problems and psychiatric disorders as a result of fears related to COVID-19 infection, the loss of family members, and financial and economic concerns due to rising unemployment and crippled economies worldwide (Marroquín, Vine, & Morgan, 2020; Rajkumar, 2020; White & Van Der Boor, 2020; Xiong et al., 2020). One population of individuals who have not received much consideration in terms of their mental health during the COVID-19 pandemic are dreamers. This group of individuals includes undocumented immigrants who entered the U.S. as minors and are eligible for Deferred Action for Childhood Arrivals (DACA), an immigration program that permits recipients to pursue an education and obtain work permits lawfully without risk of deportation. The present study was designed to gain a preliminary understanding of the mental health impacts of the COVID-19 pandemic among dreamers currently enrolled in college.

In 2017, it was estimated that about 241,000 dreamers were currently enrolled in colleges across the United States (Capps, Fix, & Jong, 2017), and that DACA recipients would contribute more than $460.3 billion to the U.S. Gross Domestic Product over the next ten years (Svajlenka, Jawetz, & Bautista-Chavez, 2017). However, despite protections from deportation, DACA recipients continue to face marginalization, socioeconomic challenges, and reduced social integration (Siemons, Raymond-Flesch, Auerswald, & Brindis, 2017), making this population vulnerable to mental health issues. The current political climate has also increased immigration concerns among dreamers, including the fear that family and friends who are not protected by DACA may be deported or detained (Siemons et al., 2017). Previous research has suggested that immigration concerns, in addition to economic and societal disadvantages, can lead to various mental health issues among DACA recipients, including anxiety and depression (Siemons el at., 2017; Enriquez, Hernandez, & Ro, 2018).

Dreamers enrolled in college potentially face a double burden as they struggle to overcome the challenges of their immigration status while also facing typical stressors related to a college education. For many students, college proves to be a stressful time. Concerns about academic performance, finances, being away from home and family, new social relationships, and plans after college are some of the factors that can have a negative impact on the mental health of college students (Aselton, 2012; Beiter et al., 2015). Visits to the campus clinic for problems related to mental health have risen steadily over the past 20 years with stress, anxiety, and depression among the top concerns (American College Health Association, 2018). Dreamers in college potentially experience the same challenges to their mental health but with added concerns about their future in the U.S. and the safety of undocumented family and friends (Teranishi, Suárez-Orozco, & Suárez-Orozco, 2015). In particular, undocumented college students face much higher rates of stress, anxiety, and depression compared to their documented peers (Potochnick & Perreira, 2010; Suárez-Orozco et al., 2015; Terriquez, 2015; O’Neal et al., 2016; Enriquez et al., 2018).

The COVID-19 pandemic has introduced new challenges that have the potential to further harm mental health. Much research has provided evidence of increased rates of stress, anxiety, and depression, among other mental health problems, as a result of the economic fallout caused by the COVID-19 pandemic (for review, see Xiong et al., 2020). College students in particular have faced a greater risk of developing mental health problems and psychiatric disorders during the pandemic (González-Sanguino et al., 2020; Lei, Huang, Zhang, Yang, Yang, & Xu, 2020), including increased anxiety and depression (Huckins et al., 2020). Moreover, in the United States, the pandemic has disproportionately impacted racial and ethnic minorities. Black and Latinx Americans experience much higher rates of COVID-19 infection and death, compared to non-Latinx Whites (Oppel Jr, Gebeloff, Lai, Wright, & Smith, 2020). In addition, unemployment rates have been consistently higher and job recovery has been slower among Black and Latinx individuals throughout the pandemic, compared to non-Latinx Whites (Sáenz & Sparks, 2020). Research also suggests that both Latinx and foreign-born individuals experience greater “fear of COVID-19” than non-Latinx and native-born individuals in the United States (Fitzpatrick, Harris, & Drawve, 2020), paving the way for greater mental health distress among these marginalized groups.

To our knowledge, no studies have empirically examined the mental health consequences of the COVID-19 pandemic for dreamers, in particular for those currently enrolled in college. In contrast to U.S. citizens, undocumented immigrants are largely ineligible for government assistance, including unemployment benefits and stimulus relief plans established for the pandemic (i.e., the CARES act). In addition, many undocumented immigrants in the United States do not receive adequate healthcare, and it has been reported that mental health is the most unmet need among this population (Raymond-Flesch, Siemons, Pourat, Jacobs, & Brindis, 2014). Recently, several investigators have hypothesized that, without appropriate economic assistance and mental healthcare, undocumented immigrants will face a disproportionate rise in mental health problems during the COVID-19 pandemic (Clark, Fredricks, Woc-Colburn, Bottazzi, & Weatherhead, 2020; Garcini, Domenech Rodríguez, Mercado, & Paris, 2020; Page, Venkataramani, Beyrer, & Polk, 2020; Wilson & Stimpson, 2020).

One of the barriers to performing research on the mental health of college dreamers is that it can be difficult to quickly find a large sample. Some schools do not allow undocumented students to enroll (regardless of DACA status), and dreamers may not reveal their immigration status for fear of being kicked out or facing discrimination from their peers. Moreover, many colleges do not collect data that would allow them to identify dreamers, or they may not release the information to investigators for research. For the present study, we recruited a readily available and fortuitous population of undocumented college students currently enrolled at a public university in Delaware. These students are DACA recipients who have been awarded a scholarship from TheDream.US.

The present study employed an online survey procedure to investigate the prevalence of mental health problems and concerns among this population of college dreamers during the COVID-19 pandemic. The study used brief clinical assessments to measure anxiety, depression, and stress, while also investigating various factors potentially related to mental health, such as academics, COVID-19 infection, immigration concerns, and unemployment. The specific purpose of the present study was three-fold, (1) to determine rates of mental health issues (i.e., anxiety, depression, and stress) and concerns among the present population of college dreamers, (2) to compare the observed proportions of mental health issues and concerns with available data from the general population of college students, and (3) to determine whether concerns regarding academics, COVID-19 infection, immigration, and unemployment (among other factors) were associated with greater clinical measures of anxiety, stress, and depression among the dreamers. Despite some limitations, the present study represents an important preliminary step in understanding the mental health consequences of the COVID-19 pandemic for this critically understudied demographic group.

## Method

### Participants and Recruitment

Participants were recruited from a population of approximately 150 undocumented undergraduate college students currently enrolled at a public university in Delaware. In total, 109 individuals from this population participated in the survey (mean age = 21.76, SD = 2.43; 73 women, 35 men, and 1 “other”). These students are DACA recipients who have been awarded a scholarship from TheDream.US. This scholarship covers out-of-state tuition and other expenses allowing dreamers from “locked-out states” (i.e., states with policies that restrict their access to college) to attend one of four partner colleges to obtain a four-year degree.

For recruitment, on May 15, 2020, an email was sent to this population of dreamers inviting them to take part in a voluntary anonymous survey “to learn more about the pandemic’s impact on dreamers and their families.” A link to the survey was provided in the email. The survey opened on May 15^th^ and closed on May 22^nd^. All participants who completed the survey were paid $15 via PayPal using an email address that the respondent provided at the end of the survey. Email addresses were not associated with an individual’s survey data and were deleted immediately upon payment.

### Measures

The survey contained questions pertaining to demographic information (age, gender, race, ethnicity, country of origin, household income, etc.), mental health, education, immigration, employment, and COVID-19 infection. The survey also contained 3 mental health assessments, including the Generalized Anxiety Disorder scale – 7 item (GAD-7), Patient Health Questionnaire – 9 item (PHQ-9), and the Perceived Stress Scale – 10 item (PSS-10), which were used as preliminary screens for anxiety, depression, and stress, respectively. For each assessment, the respondent read a list of symptoms and rated how often they experienced each symptom over the past two weeks (0 = Not at all, 1 = Several days, 2 = More than half the days, 3 = Nearly every day) for the GAD-7 and PHQ-9, and over the past month for the PSS-10 (0 = Never, 1 = Almost never, 2 = Sometimes, 3 = Fairly often, 4 = Very often). The GAD-7 and PHQ-9, in particular, are useful for predicting the presence of anxiety and depressive disorders. In the clinical setting, a score of 10 or greater on the GAD-7 or PHQ-9 serves as a cutoff point, suggesting the patient should be further evaluated for anxiety or depression, respectively. This is based on research indicating a PHQ-9 score ≥ 10 has a sensitivity of 88% and a specificity of 88% for major depression (Kroenke, Spitzer, & Williams, 2001). Likewise, a GAD-7 score ≥ 10 has a sensitivity of 89% and a specificity of 82% for generalized anxiety disorder (Spitzer, Kroenke, Williams, & Löwe, 2006). In addition, we employed the final item on the PHQ-9 (“Thoughts that you would be better off dead, or thoughts of hurting yourself in some way”) as a measure of suicidal ideation with a cutoff point of ≥ 1. In order to enhance the comfort and security of this particularly sensitive population of students, participants had the opportunity to skip survey questions. Non-responses to a survey question were excluded from percentage calculations and statistical procedures. If a participant skipped an item on a mental health assessment, their total score on the assessment was similarly excluded from all analyses and percentage calculations. In comparing our findings to other reports on college students, we employed observed vs. expected analyses (i.e., chi-square test) computed on the number of observed cases in our sample versus the number of expected cases as determined by adopting the proportions reported in these comparison studies. It should be noted that, while we present our data alongside recent findings from the U.S. general population, we only apply statistical analyses when comparing our data to recent reports on the mental health of college students. We believe that the general population of college students serves as a more fitting benchmark than the overall U.S. general population when it comes to understanding the mental health of undocumented college students in our sample, and therefore believe such comparisons are worthy of statistical analysis.

## Results and Discussion

### Participant characteristics

General demographics and characteristics about the sample of college dreamers who participated in the present study are presented in Table 1. There were 109 participants in total (mean age = 21.76, SD = 2.43; 73 women, 35 men, and 1 “other”). The majority of respondents reported having an ethnic background of Mexican, Hispanic, Latinx, or Spanish origin (94%, n = 103) and identified Mexico as their home country of origin (77%, n = 84). The participants currently resided in predominantly U.S. States of the South, including North Carolina (38%, n = 41), Georgia (32%, n = 35), and South Carolina (11%, n = 12). Most participants lived in relatively low-income households with 20% reporting an annual household income under $15,000 and 30% reporting a household income between $15,000 and $29,999. We also found that the majority of respondents had plans to pursue a graduate or professional degree after graduation (57%, n = 62). The characteristics of our sample in terms of age, ethnicity, country of origin, household income, and post-graduation plans are largely consistent with prior research on the general population of undocumented college students (Teranishi et al., 2015).

**Table 1.**

| **Participant Characteristics** |  |
| --- | --- |
| **Total Number of Participants** | 109 |
| **Age (Mean ± SD)** | 21.76 ± 2.43 |
| **Sex (n)** |  |
| Female | 66.97% (73) |
| Male | 32.11% (35) |
| Other | 1% (1) |
| **Race (n)** |  |
| American Indian / Alaskan Native | 2.75% (3) |
| Asian | 1.83% (2) |
| Black / African American | 5.50% (6) |
| Native Hawaiian / Pacific Islander | 0.92% (1) |
| Two or More Races | 3.67% (4) |
| White | 50.46% (55) |
| Other | 29.36% (32) |
| Not Answered | 5.50% (6) |
| **Ethnicity (n)** |  |
| Mexican, Mexican American, Chicano | 76.15% (83) |
| Another Hispanic, Latino, or Spanish origin | 18.35% (20) |
| Not of Hispanic, Latino, or Spanish origin | 5.50% (6) |
| **Home Country (n)** |  |
| Mexico | 77.06% (84) |
| Honduras | 7.34% (8) |
| El Salvador | 4.59% (5) |
| Other | 11.00% (12) |
| **Current State Residence (n)** |  |
| North Carolina | 37.61% (41) |
| Georgia | 32.11% (35) |
| South Carolina | 11.01% (12) |
| Delaware | 6.42% (7) |
| Other | 12.85% (14) |
| **Gross Household Income for 2019 (n)** |  |
| Under \$15,000 | 20.18% (22) |
| Between \$15,000 and \$29,999 | 29.36% (32) |
| Between \$30,000 and \$49,999 | 18.35% (20) |
| Between \$50,000 and \$74,999 | 2.75% (3) |
| Between \$75,000 and \$99,999 | 2.75% (3) |
| Unknown | 26.61% (29) |
| **What are your plans after graduation? (n)** |  |
| Pursue Graduate or Professional Degree | 56.88% (62) |
| Work | 40.37% (44) |
| Other | 2.75% (3) |

### Mental Health Assessments: Anxiety, Depression, and Stress

#### Results

The present survey included mental health assessments for anxiety and depression (the GAD-7 and PHQ-9, respectively) in addition to a subclinical scale measuring stress (PSS-10). The results are presented in Table 2 and Figure 1. In our sample, 47% of the respondents met the clinical cutoff for anxiety (GAD-7 score ≥ 10), 63% met the cutoff for depression (PHQ-9 ≥ 10), and 67% met the cutoff for anxiety and/or depression. Moreover, 29% of the respondents met the cutoff for suicidal ideation. The mean score for the PSS-10 was 23.26 ± 6.30. The present findings suggest a high proportion of undocumented college students may be experiencing moderate-to-severe anxiety, depression, suicidal ideation, and stress during the COVID-19 pandemic. Furthermore, it is likely most respondents in our sample would meet the criteria for a full diagnosis of either an anxiety or depressive disorder upon further evaluation. There were no sex differences observed in any of the mental health measures.

**Table 2.**
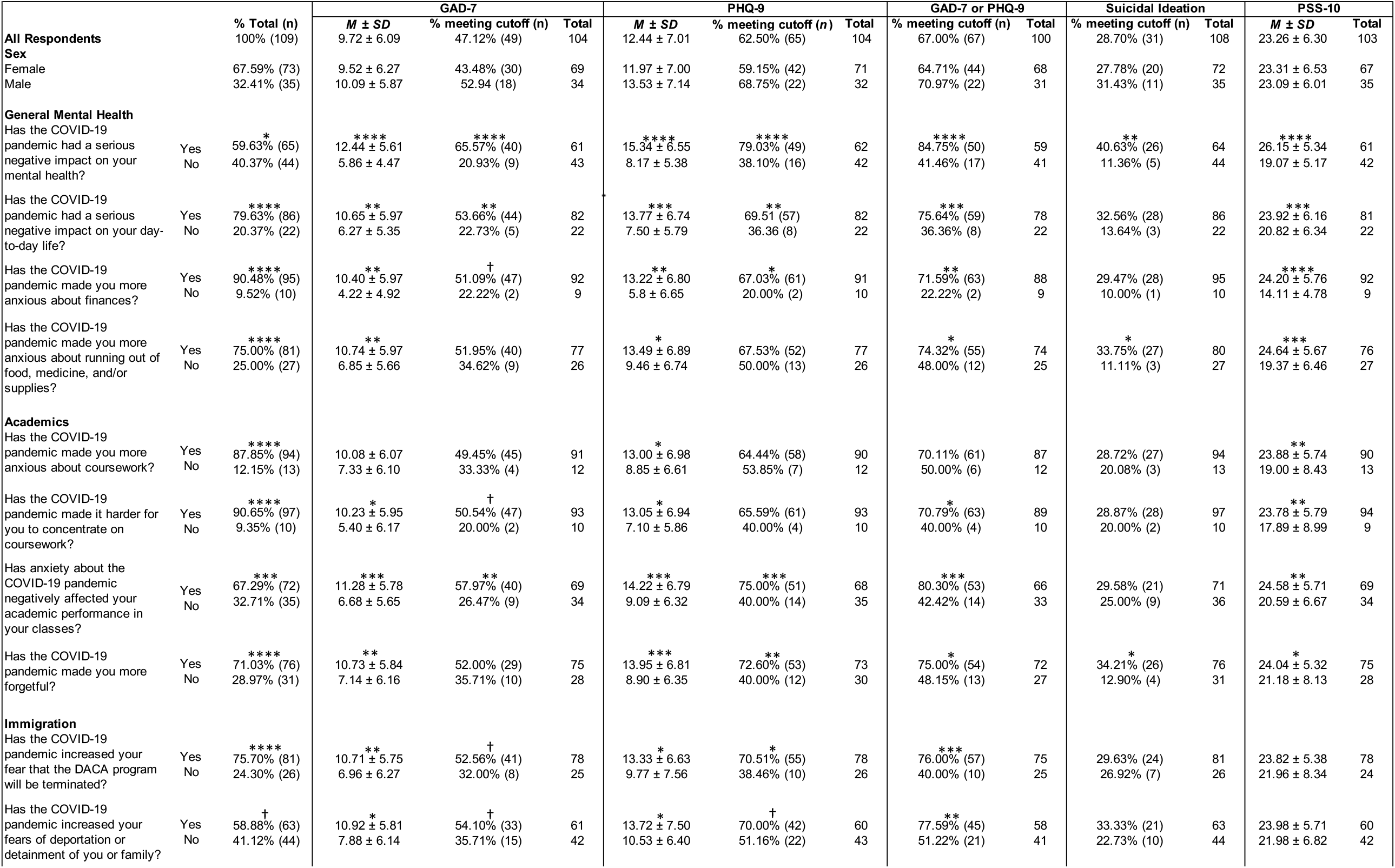
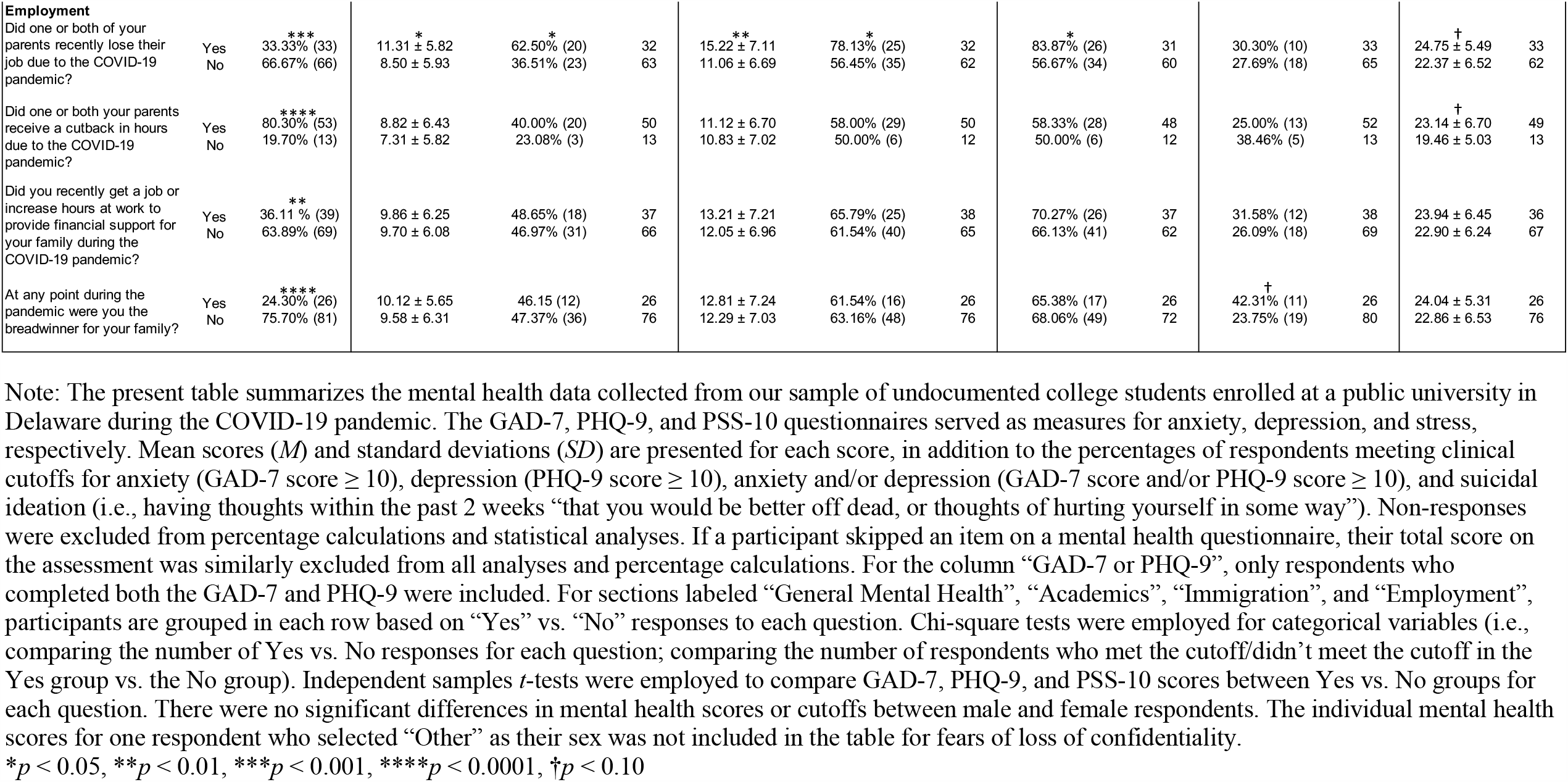
Mental health of undocumented college students during the COVID-19 pandemic

**Figure 1.**
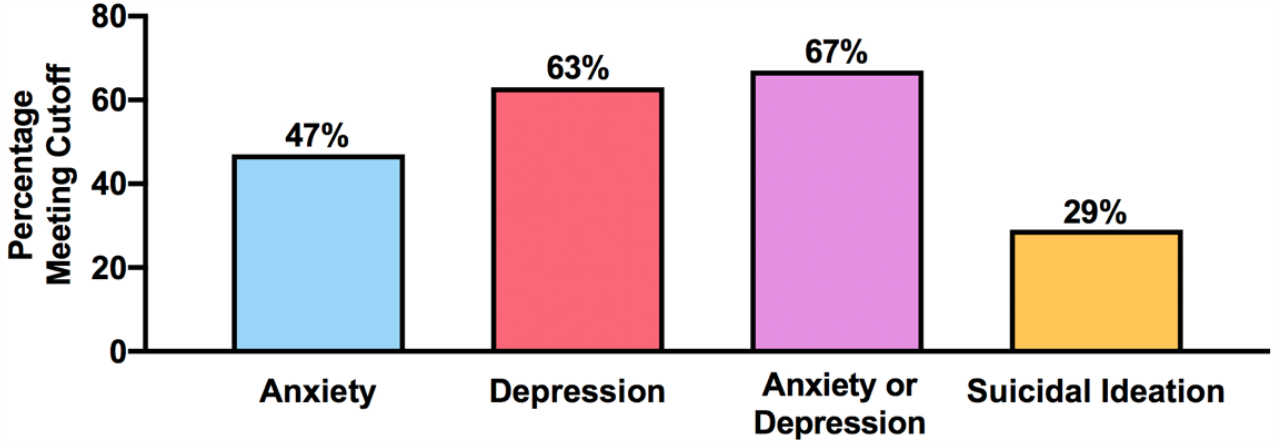
Percentages of undocumented college students in our sample meeting clinical cutoffs for anxiety (GAD-7 score ≥ 10), depression (PHQ-9 score ≥ 10), anxiety or depression (GAD-7 score and/or PHQ-9 score ≥ 10), and suicidal ideation (i.e., having thoughts within the past 2 weeks “that you would be better off dead, or thoughts of hurting yourself in some way”). Findings suggest a high proportion of undocumented college students are suffering from anxiety and depressive symptoms, in addition to suicidal ideation, during the COVID-19 pandemic. Respondents would likely meet criteria for a full diagnosis of an anxiety or depressive disorder upon further evaluation.

#### Comparisons with prior research on undocumented college students

The rate of anxiety observed in the present study was higher than previously reported for college dreamers (see Figure 2). A national survey conducted as part of the UndocuScholars Project in 2015 found that 35% of college dreamers met the clinical cutoff for anxiety based on the GAD-7 (Teranishi et al., 2015), compared to 47% in our sample. To determine whether the difference between studies was significant, we performed a chi-square test comparing our observed outcome (47% of our sample meeting the cutoff for anxiety [n = 49] and 53% not meeting the cutoff [n = 55]) versus an expected outcome based on Teranishi et al., 2015 (35% of the sample meeting the cutoff [n = 36] and 65% not meeting the cutoff [n = 68]). This analysis revealed a significant difference between observed versus expected outcomes (χ^2^ = 7.18, *p* = 0.007), suggesting a higher rate of anxiety in the present sample of college dreamers compared to the expected outcome based on the 2015 report. The higher rate of anxiety in our study may be attributed in part to the COVID-19 pandemic. However, we cannot rule out the possibility of other variables being involved, such as general immigration concerns which have been increasing for the past several years (Kirsten & Boneparth, 2017; García, 2018). In addition, the 2015 study involved a national survey of college dreamers, whereas the present sample of college dreamers all attended the same university and mainly resided in the Southern U.S. Therefore, it is possible that differences in the demographic characteristics of each sample may have partially contributed to the observed differences in mental health. However, in contrast to this alternative explanation, the present sample shared important demographic characteristics with college dreamers in the 2015 study, including similar means and proportions of age, ethnicity, country of origin, household income, and post-graduation plans (Teranishi et al., 2015).

**Figure 2.**
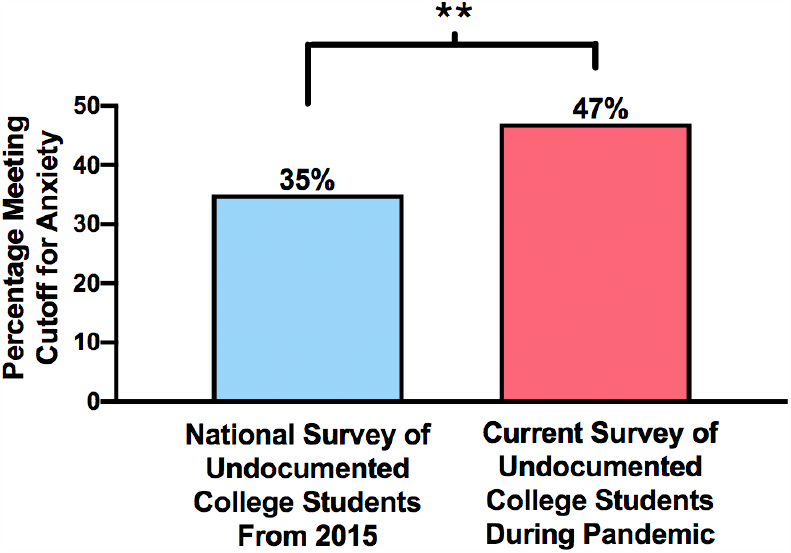
Percentage of undocumented college students meeting cutoff for anxiety (GAD-7 score ≥ 10) in 2015 UndocuScholars survey (Teranishi et al., 2015) versus the present survey conducted during the COVID-19 pandemic. Observed vs. expected analysis revealed a significantly higher number of undocumented college students in the present survey met the cutoff for anxiety, compared to an expected number based on the 2015 UndocuScholars survey. **p < 0.01

The dreamers in our sample also showed a higher depression score compared to a previous study on the mental health of undocumented college students (O’Neal et al., 2016). In this previous study, investigators examined depression in a sample of 84 first-generation Latinx “non-citizen” college students (O’Neal et al., 2016). Approximately 60% of this sample was undocumented. The mean PHQ-9 score in this previous study was 8.83 ± 6.80, which is significantly lower than the depression score in our study of college dreamers (*t*(186) = 3.56, *p* = 0.0005). In another recent study, depression was examined in a small sample of undocumented college students (n = 12) with a mean PHQ-9 score of 7.33 ± 6.05 (Campos, 2020). This figure was also significantly lower than the depression score in our sample of dreamers (*t*(114) = 2.42, *p* = 0.017).

In addition to anxiety and depression, we observed a high level of perceived stress in our sample, based on the results from the PSS-10. Unlike the GAD-7 and PHQ-9, the PSS-10 is not employed as a diagnostic tool, and no cutoff points have been officially established for this scale. However, it is worth noting that an early study using the PSS-10 reported a mean and standard deviation of 13.02 ± 6.35 for the general U.S. population and 14.2 ± 6.2 for U.S. respondents between the ages of 18–29 (Cohen & Williamson, 1988), and our current study of college dreamers produced a much higher figure for perceived stress (23.21 ± 6.29). Notwithstanding a high level of perceived stress in our sample, it is not clear whether perceived stress in college dreamers had actually changed as a result of the pandemic. Indeed, previous research suggests that perceived stress among undocumented college students is already very high. One study reported an average PSS-10 score of 22.52 ± 4.99 in a sample of first-generation Latinx “non-citizen” college students (O’Neal et al., 2016), comparable to the perceived stress level in our sample of college dreamers (*t*(185) = 0.871, *p* = 0.385).

#### Comparisons with the general population of college students during the COVID-19 pandemic

Importantly, the rate of respondents meeting the clinical cutoffs for anxiety and depression in the present study were higher than those recently reported for college students overall (Figure 3). A large national survey of college students was recently conducted by the Healthy Minds Network (HMN) and American College Health Association (ACHA) to learn more about the mental health of college students during the COVID-19 pandemic. In this national survey, 31% of college students met the clinical cutoff for anxiety and 41% met the cutoff for depression, based on responses to the GAD-7 and PHQ-9, respectively (Healthy Minds Network and American College Health Association, 2020). To determine whether these proportions were significantly different from our study, we performed a chi-square test comparing our observed outcomes for anxiety and depression versus what would be expected based on the HMN/ACHA survey. Our observed proportion of college dreamers meeting the cutoff for anxiety (47% [n = 49] met the cutoff; 53% [n = 55] did not meet the cutoff) was significantly higher than expected based on the proportion of respondents with anxiety in the HMN/ACHA survey (31% [n = 32] meeting the cutoff; 69% [n = 72] not meeting the cutoff), χ^2^ = 13.05, *p* = 0.0003. In addition, our observed proportion of college dreamers meeting the cutoff for depression (62.5% [n = 65] met the cutoff; 37.5% [n = 39] did not meet the cutoff) was also higher than expected based on the HMN/ACHA findings (41% [n = 43] meeting the cutoff; 59% [n = 61] not meeting the cutoff), χ^2^ = 19.19, *p* < 0.0001. The findings suggest that the undocumented college students in our survey demonstrated significantly higher proportions of anxiety and depression during the COVID-19 pandemic, compared to college students overall.

**Figure 3.**
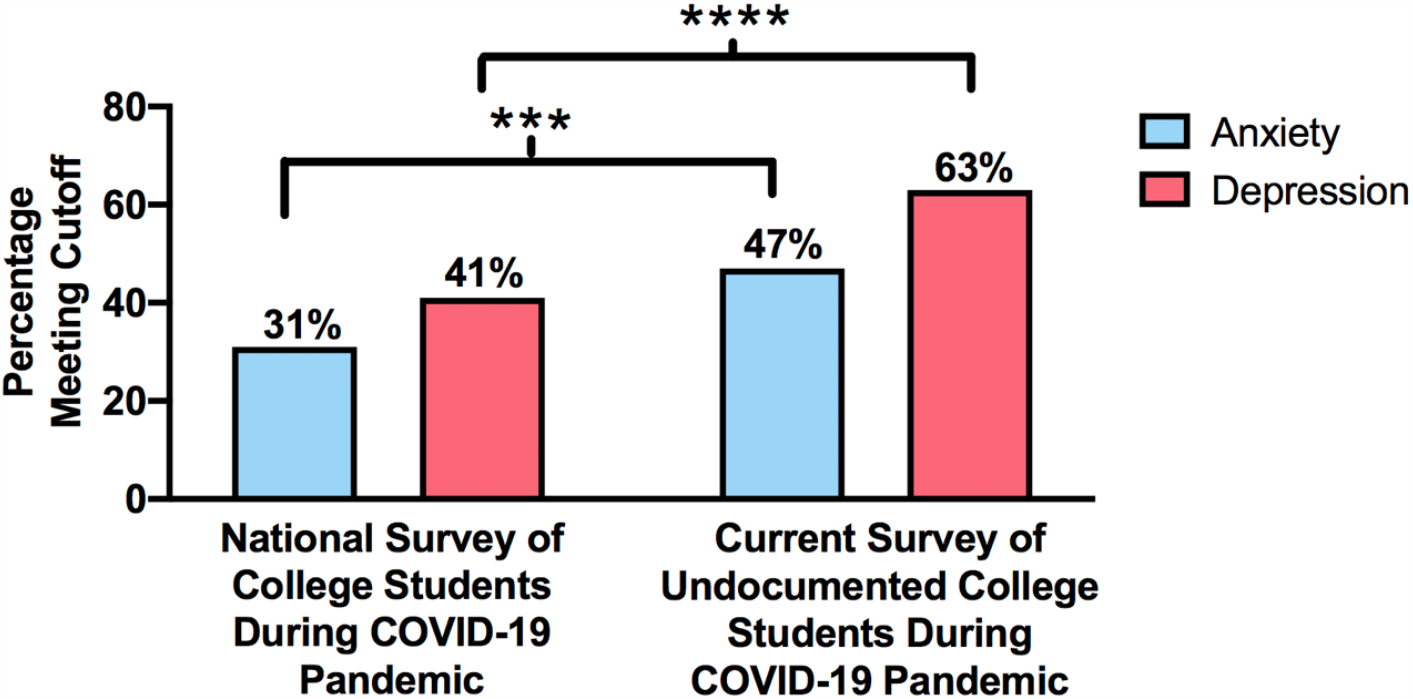
Percentages of college students meeting clinical cutoffs for anxiety and depression in a national survey, compared to undocumented college students in the present survey, during the COVID-19 pandemic. The national survey of college students was conducted by the Healthy Minds Network and the American College Health Association (HMN/ACHA). The GAD-7 and PHQ-9 were employed to measure anxiety and depression, respectively. The cutoff for each scale was a score ≥ 10. Observed vs. expected analyses revealed a significantly higher number of undocumented college students met the cutoffs for anxiety and depression, compared to expected outcomes based on the HMN/ACHA national survey of college students. \*\*\**p* < 0.001, \*\*\*\**p* < 0.0001

#### Comparisons with the overall U.S. general population during the COVID-19 pandemic

The proportions of respondents meeting the clinical cutoffs for anxiety and depression in our sample were also higher than measures recently taken from the U.S. general population during the pandemic (Figure 4). The Centers for Disease Control and Prevention (CDC) recently conducted a national survey of Americans (i.e. the Household Pulse Survey) to measure changes in mental health during the COVID-19 pandemic. The survey began on April 23, 2020 and continued for several months, providing weekly national measures of anxiety and depression using the GAD-2 and PHQ-2, respectively. According to CDC data from the week of May 14– 19^th^ (which overlapped the time frame of our study), 28% of Americans across the U.S. met the clinical cutoff for anxiety, 24% met the clinical cutoff for depression, and 34% met the clinical cutoff for anxiety and/or depression. These national figures are considerably lower than the high rates of mental health issues we observed in our study of college dreamers (using the GAD-2 and PHQ-2, the percentages in our study were 45%, 50%, and 64%, respectively; see Figure 4). Notably, the rate of depression among our sample of college dreamers doubled the national rate (50% vs. 24%). It should be mentioned, however, that certain factors mitigated these differences, such as age and ethnicity. In the CDC study, higher rates of anxiety and depression were observed for respondents in the 18–29 age group (39%, 37%, and 48%) and for Hispanic/Latino respondents (34%, 29%, and 41%). However, the college dreamers in our study continued to display higher rates of anxiety and depression, even when compared to these individual groups in the CDC survey.

**Figure 4.**
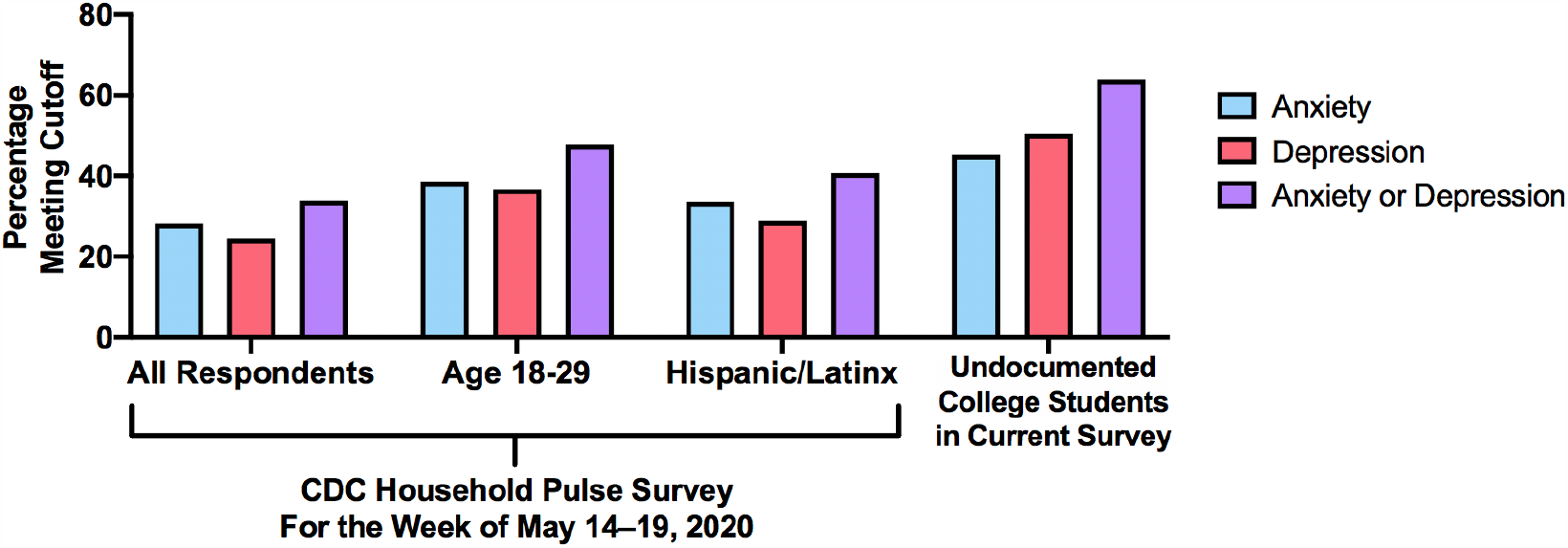
Percentages of national survey respondents meeting cutoffs for anxiety, depression, and anxiety and/or depression in the CDC Household Pulse Survey, compared to undocumented college students in the present survey, during the COVID-19 pandemic. Anxiety and depression were measured using GAD-2 and PHQ-2, respectively, instead of the GAD-7 and PHQ-9. This resulted in slightly different percentages of our sample meeting cutoffs for anxiety and depression. The cutoff for the GAD-2 and PHQ-2 was a score ≥ 3.

In contrast to emerging research on anxiety and depression, there are few studies that have focused on perceived stress during the COVID-19 pandemic, which prevents us from comparing our findings to the general population. There are preliminary reports, however, of global and national levels of perceived stress, which have been shared online, but have yet to receive peer-review (Kowal et al., 2020, preprint; Limcaoco, Mateos, Fernández, & Roncero, 2020, preprint). For instance, in a recent global survey (Limcaoco et al., 2020, preprint), average perceived stress level during the pandemic was 17.4 ± 6.4 for all respondents, 20.7 ± 7.0 for students, and 16.9 ± 5.2 for respondents in North America. As a preliminary analysis for understanding how perceived stress in our sample may compare with the general population during the pandemic, we ran independent samples *t*-tests to compare our findings on perceived stress with the global and national figures reported in Limcaoco et al. (2020). We found that the perceived stress level in our sample was significantly higher than those reported globally (*t*(1192) = 8.89, *p* < 0.0001), for students (*t*(174) = 2.53, *p* = 0.0121), and for respondents in North America (*t*(140) = 5.62, *p* < 0.0001). These findings suggest greater perceived stress in our sample of college dreamers. It should be emphasized, however, that there are critical caveats to comparing results between the two surveys. In particular, the study by Limcaoco et al. (2020) was a preliminary, unpublished report, and the survey was performed about 2 months earlier than the present survey. Thus, discrepancies in methodology and timing could partially explain the difference in perceived stress between our dreamers and the global/national levels reported in Limcaoco et al. (2020).

#### Discussion

In sum, our findings suggest that college dreamers are experiencing a range of mental health issues during the COVID-19 pandemic, including anxiety, depression, suicidal ideation, and stress. Notably, 2 in 3 college dreamers met the clinical cutoff for anxiety and/or depression. The rate of depression, in particular, doubled the national rate reported for the U.S. general population at a similar time during the pandemic (CDC, 2020). Based on preliminary analyses, the college dreamers in our sample also showed greater anxiety and depression symptoms, relative to past mental health data of college dreamers collected *before* the pandemic (Teranishi et al., 2015; O’Neal et al., 2016; Campos, 2020). While we have noted some caveats to comparing our findings with previous research on dreamers, it is tempting to speculate that our preliminary analyses represent an *increase* in depression and anxiety symptoms among college dreamers as a result of the COVID-19 pandemic. This interpretation of the data is especially reasonable considering that depression and anxiety have risen for most other demographic groups during the pandemic (CDC, 2020), including college students (Healthy Minds Network and American College Health Association, 2020). However, our findings suggest that the pandemic may be affecting the mental health of college dreamers to a more severe degree than the U.S. general population, especially when compared to college students overall.

### General Mental Health Questions

#### Results

The present survey included general questions pertaining to participants’ mental health which are presented in Table 2. We employed chi-square analyses to compare the proportion of Yes vs. No responses for each item. Missing values were excluded from the analyses. Results revealed that a significant majority of respondents (60%, n = 65) believed the COVID-19 pandemic had a serious negative impact on their mental health (χ^2^ = 4.05, *p* = 0.0443), and 80% (n = 86) reported that the COVID-19 pandemic had a serious negative impact on their day-to-day lives (χ^2^ = 37.93, *p* < 0.0001). Notably, 90% of respondents (n = 95) reported that the pandemic made them more anxious about finances (χ^2^ = 68.76, *p* < 0.0001), and 76% (n = 81) that the pandemic made them more anxious about running out of food, medicine, and/or supplies (χ^2^ = 28.27, *p* < 0.0001). There were no sex differences observed in any of these measures. Overall, the findings suggest that the vast majority of undocumented college students in our sample perceived a negative impact on their mental health as a result of the COVID-19 pandemic, especially in terms of aggravated financial concerns. Importantly, these concerns were related to greater scores on the anxiety, depression, and stress scales (as determined by independent samples *t*-tests; see Table 2).

#### Comparisons with the general population of college students during the COVID-19 pandemic

The dreamers in our sample demonstrated greater concerns over finances when compared to the general population of college students in the U.S. during the pandemic. A large national survey of 18,764 college students performed from late March through May 2020 found that 66% of respondents were experiencing financial stress due to the pandemic (Healthy Minds Network and American College Health Association, 2020). This figure was much lower than the proportion of dreamers in our sample experiencing heightened anxiety about finances due to the pandemic (90%). To determine whether the difference between studies was significant, we performed a chi-square test comparing our observed outcome (90% [n = 95] reported increased financial anxiety; 10% [n = 10] reported no increase) to an expected outcome based on the proportion of respondents reporting increased financial stress in the HMN/ACHA survey (66% [n = 69] reporting financial stress; 34% [n = 36] reporting no stress). Results indicated our observed outcome was significantly different than the HMN/ACHA expected outcome, χ^2^ = 28.57, *p* < 0.0001, suggesting a higher proportion of financial concerns in the present sample of college dreamers compared to college students overall. However, it should be noted that our survey asked participants if the pandemic made them “more anxious” about finances, while the HMN/ACHA survey asked them if their financial situation had become “more stressful” during the pandemic. We cannot rule out the possibility that differences in phrasing may be partially responsible for the difference between the two surveys.

#### Comparisons with the U.S. general population during the COVID-19 pandemic

The proportions of mental health concerns in our sample of college dreamers were also much higher than what has been observed in surveys of the U.S. general population during the COVID-19 pandemic. A poll from the American Psychiatric Association (APA) conducted in early March 2020 found that 36% of U.S. adults believed that the pandemic had a serious impact on their mental health, and 59% believed the pandemic had a serious impact on their day-to-day lives, which are lower figures than those observed in our present sample of dreamers (60% and 80%, respectively). In addition, the APA poll found that 57% of Americans were concerned about the impact of the pandemic on their finances (compared to 90% in our sample), and about half were worried about running out of food, medicine, and/or supplies (compared to 76% in our sample).

#### Discussion

Taken together, a significant majority of the college dreamers in our survey perceived an increase in mental health distress, changes to their day-to-day lives, and anxiety over finances as well as food, medicine, and/or supplies due to the COVID-19 pandemic. These concerns were related to significantly greater anxiety, depression, and stress, as measured by the GAD-7, PHQ-9, and PSS-10. The proportions of respondents expressing these concerns were higher in our sample than those reported for the U.S. general population. Moreover, while finances have proven a major concern for most Americans during the pandemic, there were more dreamers in our sample expressing financial concerns than reported in a recent national survey of the general population of college students. It should be noted that there are several reasons to be cautious when comparing the present findings with the results of other surveys. In particular, the APA survey was conducted 2 months prior to the present survey. Moreover, the items being compared across surveys contain subtle differences in phrasing. We cannot rule out the possibility that differences between those prior studies and the present findings may be attributed in part to discrepancies in how and when the surveys were performed.

### Academics, immigration, and COVID-19 concerns

#### Results

Chi-square analyses were employed to compare the proportion of Yes vs. No responses for questions related academics and immigration concerns. Missing values were excluded from the analyses. A high proportion of dreamers (88%, n = 94) reported that the pandemic made them more anxious about coursework (χ^2^ = 61.31, *p* < 0.0001), and 91% of the respondents (n = 97) had a harder time concentrating on their coursework (χ^2^ = 70.74, *p* < 0.0001). Moreover, 67% of respondents (n = 72) reported that anxiety about the pandemic hurt their academic performance (e.g., in terms of grades, completing assignments on time, etc.; χ^2^ = 12.79, *p* = 0.0003), and 71% (n = 76) said the pandemic made them more forgetful (χ^2^ = 18.93, *p* < 0.0001). “Yes” responses to all academic questions were associated with significantly greater scores of anxiety, depression, and stress scores, as determined by the GAD-7, PHQ-9, and PSS-10 scales (see Table 2). When asked how worried they were that the pandemic will prevent them from achieving their academic and professional goals (on a scale of 1 to 7; 1 = Not at All Worried and 7 = Extremely Worried), dreamers responded with a mean score of 5.24 ± 1.77, with 37% (n = 40) saying they were “extremely worried” (Table 3 and Figure 6). Worry about achieving academic and professional goals was correlated with anxiety (*r* = 0.434, *p* < 0.0001), depression (*r* = 0.436, *p* < 0.0001), and stress scores (*r* = 0.397, *p* < 0.0001; see Table 3).

**Table 3.** Correlations between survey items on Likert scales and mental health assessments for anxiety (GAD-7), depression (PHQ-9), and stress (PSS-10).

| Question | $M \pm SD$ | GAD-7 | | | PHQ-9 | | | PSS-10 | | |
| --- | --- | --- | --- | --- | --- | --- | --- | --- | --- | --- |
| | | $r$ | $p$ | CI | $r$ | $p$ | CI | $r$ | $p$ | CI |
| How worried are you that the COVID-19 pandemic will prevent you from achieving your academic and professional goals? | $5.24 \pm 1.77$ | 0.434 | 0.0000 | 0.2630 to 0.5780 | 0.436 | 0.0000 | 0.2656 to 0.5798 | 0.397 | 0.0000 | 0.2199 to 0.5481 |
| Does the possibility of the DACA program ending make you anxious? | $6.08 \pm 1.43$ | 0.204 | 0.0378 | 0.0118 to 0.3816 | 0.168 | 0.0908 | -0.0271 to 0.3513 | 0.144 | 0.1458 | -0.0506 to 0.3287 |
| How worried are you that you or an immediate family member will contract COVID-19? | $4.73 \pm 1.77$ | 0.281 | 0.0038 | 0.0939 to 0.4496 | 0.262 | 0.0073 | 0.0726 to 0.4323 | 0.191 | 0.0538 | -0.0030 to 0.3705 |
| How worried would you be about seeking treatment or having a family member seek treatment for COVID-19 due to fears of detainment or deportation? | $4.46 \pm 1.96$ | 0.347 | 0.0003 | 0.1656 to 0.5059 | 0.272 | 0.0053 | 0.0834 to 0.4412 | 0.334 | 0.0006 | 0.1498 to 0.4952 |
| Overall, how anxious are you about the COVID-19 pandemic? | $5.12 \pm 1.44$ | 0.422 | 0.0000 | 0.2485 to 0.5688 | 0.424 | 0.0000 | 0.2492 to 0.5720 | 0.386 | 0.0001 | 0.2068 to 0.5398 |

Note: Responses to all items were recorded on a 7-point Likert scale with 1 = Not At All Worried and 7 = Extremely Worried, or 1 = Not At All Anxious and 7 = Extremely Anxious. $p < 0.05$ indicates a statistically significant correlation.

**Figure 5.**
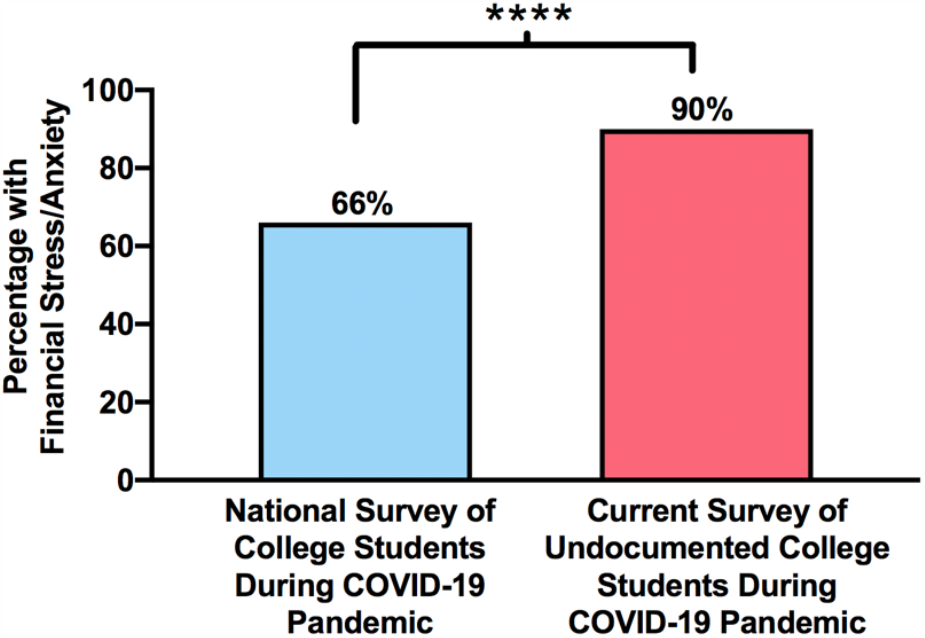
Percentage of national college students reporting the pandemic has made their financial situation “more stressful” vs. undocumented college students in our sample reporting the pandemic has made them “more anxious” about finances. The national survey of college students was conducted by the Healthy Minds Network and the American College Health Association (HMN/ACHA). Observed vs. expected analysis revealed a significantly higher number of undocumented college students reported increased financial anxiety, compared to expected outcomes based on the HMN/ACHA national survey of college students. \*\*\*\**p* < 0.0001

**Figure 6.**
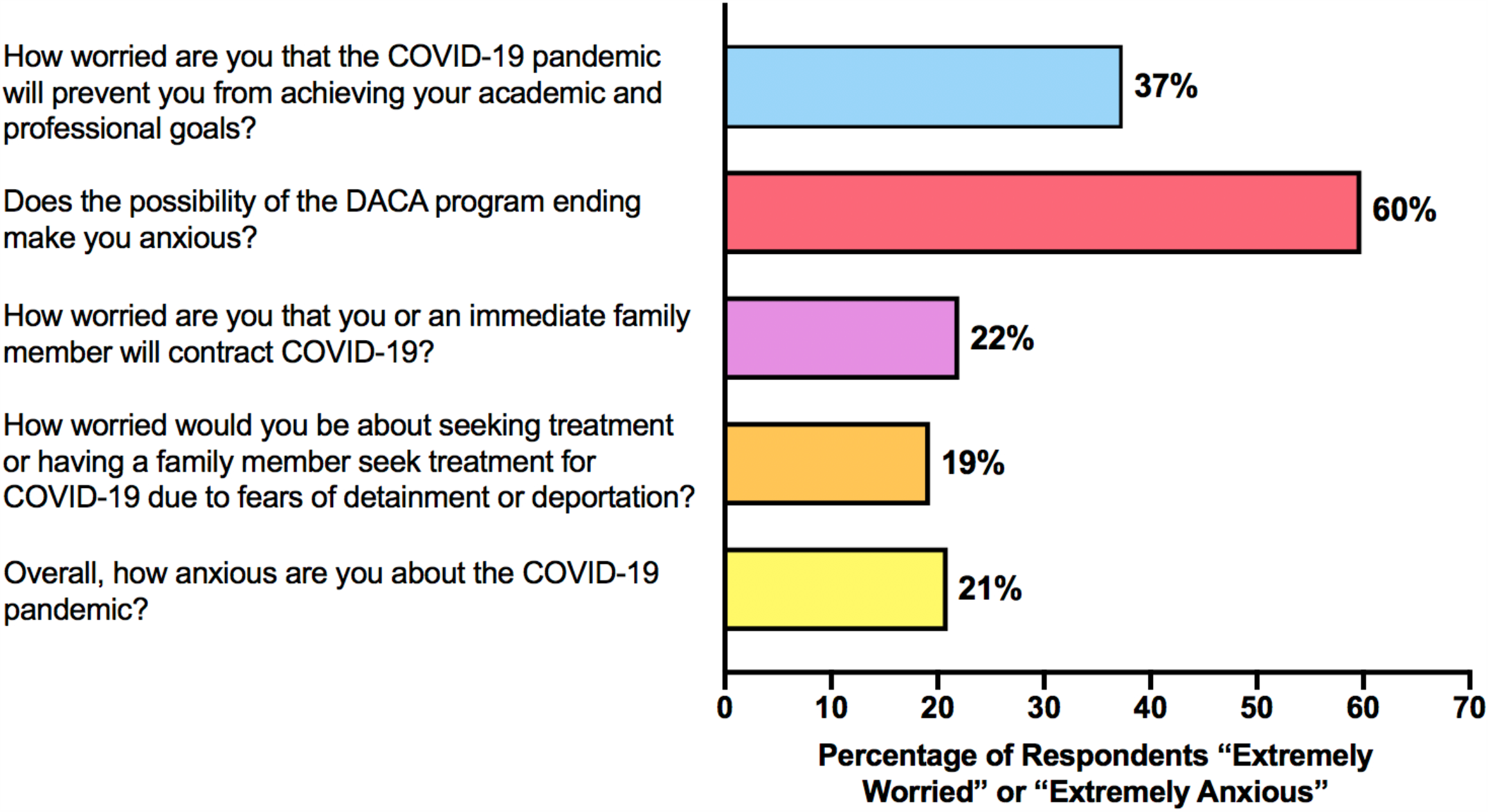
Percentages of undocumented college students in our sample who reported they were “Extremely Worried” or “Extremely Anxious” for each question. Responses were recorded on a 7-point Likert scale with 1 = Not At All Worried and 7 = Extremely Worried, or 1 = Not At All Anxious and 7 = Extremely Anxious.

Regarding how the COVID-19 pandemic affected immigration concerns, 76% of respondents (n = 81) indicated that the pandemic increased their fears that the DACA program will be terminated (χ^2^ = 28.27, *p* < 0.0001), and 59% (n = 63) reported that the pandemic increased their fears of deportation or detainment of themselves or their families (χ^2^ = 3.75, *p* = 0.0662). Heightened fear about possible DACA termination was associated with greater anxiety (*t*(101) = 2.77, *p* = 0.007) and depression (*t*(102) = 2.290, *p* = 0.0241), but not stress (t(100) = 1.288, *p* = 0.201), according to GAD-7, PHQ-9, and PSS-10 scores. Moreover, when asked how anxious they were about the possibility of the DACA program ending (on a scale of 1 to 7; 1 = Not at All Anxious and 7 = Extremely Anxious), respondents yielded a mean score of 6.08 ± 1.43, with 60% saying the possibility made them “extremely anxious.” Responses to this item were also correlated with GAD-7 anxiety score (*r* = .204, *p* = 0.038).

Questions about COVID-19 infection are presented in Table 4. In general, rates of COVID-19 infection were low. None of the respondents reported testing positive for COVID-19, and only a few respondents indicated that a member of their immediately family tested positive.

**Table 4.** Questions about COVID-19 infection

| Question | % Yes (n) | % No (n) | Total |
| --- | --- | --- | --- |
| Have you tested positive for COVID-19? | 0% (0) | 100% (109) | 109 |
| Has anyone in your immediate family tested positive for COVID-19? | 5.50% (6) | 94.50% (103) | 109 |
| • If yes, do you feel they were able to receive adequate medical care and treatment? | 33.33% (2) | 66.67% (4) | 6 |
| Has anyone in your immediate family died from COVID-19? | 0.92% (1) | 99.08% (108) | 109 |
| At any time, did you suspect you or a family member had COVID-19 but did not get tested for fear of detainment or deportation? | 10.09% (11) | 89.91% (98) | 109 |
| At any time, did you or a family member seek a test for COVID-19 but were denied? | 3.67% (4) | 96.33% (105) | 109 |
| Does everyone in your immediate family have health insurance? | 7.34% (8) | 92.66% (101) | 109 |

As expected, the vast majority of respondents (93%, n = 101) reported that at least one member of their immediate family did not currently have health insurance. When asked how worried they were that they or an immediate family member would contract COVID-19, 22% of respondents said they were “extremely worried” (Figure 6) and this was correlated with anxiety (*r* = 0.281, *p* = 0.0038) and depression (*r* = 0.262, *p* = 0.0073; Table 3). Moreover, 19% of respondents said they would be “extremely worried” about seeking treatment or having a family member seek treatment for COVID-19 due to fears of detainment or deportation. Worry about seeking treatment was correlated with anxiety (*r* = 0.347, *p* = 0.0003), depression (*r* = 0.272, *p* = 0.0053), and stress (*r* = 0.334, *p* = 0.0006). Finally, when asked how anxious they were about the COVID-19 pandemic overall, 21% reported being “extremely anxious”, and these scores were similarly correlated with anxiety (*r* = 0.442, *p* < 0.0001), depression (*r* = 0.424, *p* < 0.0001), and stress (*r* = 0.386, *p* = 0.0001).

#### Discussion

Taken together, the findings indicate that college dreamers are experiencing increased anxiety about a variety of factors as a result of the COVID-19 pandemic, including coursework, achieving future academic and professional goals, DACA termination, detainment/deportation, and COVID-19 infection. Importantly, each of these concerns was associated with a significantly higher mental health score on the GAD-7, PHQ-9, and/or PSS-10. The findings are consistent with the hypothesis that increased concerns about academics, immigration, and health as a result of the COVID-19 pandemic might be related to clinically higher levels of anxiety, depression, and stress in college dreamers. However, one question that remains undetermined is whether the association between these pandemic-related concerns and measures of mental health represents a *causal* relationship. Moreover, the present findings do not indicate the direction of the relationship. For instance, while it possible that heightened anxiety about immigration during the COVID-19 pandemic caused an appreciable increase in generalized anxiety (as measured by the GAD-7), an alternative explanation is that those with higher anxiety may be more prone to an increase in immigration concerns during the pandemic. Further research will be required to determine the direction of the relationship, in addition to causality.

In terms of academics, it is noteworthy that reports of worse academic performance, trouble concentrating on coursework, and forgetfulness were all related to a higher anxiety score on the GAD-7. Several investigators have recently suggested that heightened anxiety during the COVID-19 pandemic might impair cognitive processes, such as memory and concentration (Boals & Banks, 2020; Ritchie, Chan, & Watermeyer, 2020). It is well known that anxiety impairs memory and other executive functions, and that these effects are mediated, in part, by the actions of stress hormones in the hippocampus and prefrontal cortex (Goodman, Packard, and McIntyre, 2017). Therefore, it is possible that, in the present study, increased anxieties related to the COVID-19 pandemic might have produced cognitive impairments and, consequently, worse academic performance in our sample of college dreamers. While some anecdotal reports of forgetfulness during the pandemic have been published (e.g., Purtill, 2020), the present survey is the first to our knowledge to include a question about forgetfulness caused by the pandemic. Indeed, a significant majority of the college dreamers in our sample (71%) reported an increase in forgetfulness as a result of the pandemic, and this group also demonstrated higher anxiety, stress, and depression scores, consistent with the hypothesized negative impact of mental health on memory during the pandemic (Boals & Banks, 2020; Ritchie et al., 2020).

Interestingly, the pandemic putatively *increased* immigration concerns in a majority of the college dreamers in our sample. The reason for this remains unknown, as there is not a clear link between the pandemic and the potential for DACA termination and detainment/deportation. In fact, at the time the survey was given, the U.S. had reportedly halted detainments and deportations of undocumented immigrants (but see also Miroff, 2020). One potential explanation for the increase in immigration-related concerns is fear generalization (Dunsmoor & Paz, 2015). That is, the pandemic may have produced a heightened state of fear and anxiety that generalized across multiple domains, including immigration concerns. It should be noted that the present survey was conducted in May 2020, and that the U.S. Supreme Court had planned to rule on the constitutionality of DACA in June 2020. As a result, most of the dreamers in our sample reported they were “extremely anxious” about the possibility of the DACA program ending. On June 18, 2020, the Supreme Court ruled to uphold DACA, but at the same time the Court provided a strategy that the presidential administration could employ to terminate DACA lawfully. Moreover, at the time of this writing, U.S. Citizenship and Immigration Services has continued to deny new DACA applications. Therefore, it is not clear whether the June 2020 Supreme Court decision did anything to allay dreamers’ concerns about the possibility of DACA termination.

Finally, while reports of COVID-19 infections among respondents and their family members remained low, about 1 in 5 respondents reported that they were “extremely worried” that they or an immediately family member might contract COVID-19. In addition, 1 in 5 respondents reported they would be “extremely worried” about seeking treatment or having a family member seek treatment for COVID-19 due to fears of detainment or deportation. These findings are consistent with predictions about how the COVID-19 pandemic might disproportionately impact the mental health and wellbeing of undocumented immigrants (Clark et al., 2020; Garcini et al., 2020; Page et al., 2020; Wilson & Stimpson, 2020). Even though undocumented immigrants are at greater risk of pandemic-related health issues (including not only COVID-19 infection but also mental health distress), prior evidence suggests that this population might be less likely to seek treatment due, in part, to concerns about deportation (Maldonado, Rodriguez, Torres, Flores, & Lovato, 2013; Hacker, Anies, Folb, & Zallman, 2015), which is consistent with the present findings.

### Employment

#### Results

The findings from survey items related to employment and unemployment are presented in Table 2. Chi-square analyses were employed to compare the proportion of Yes vs. No responses. Missing values were excluded from the analyses. Among the present sample of college dreamers, 33% reported that one or both parents recently lost their job due to the COVID-19 pandemic (χ^2^ = 11.00, *p* = 0.0009; respondents who reported living with neither parent [n = 8] were excluded from this analysis). Among the respondents who reported no parental job loss, a significant majority (80%) reported that at least one parent received a cutback in hours due to the pandemic (χ^2^ = 24.24, *p* < 0.0001). Thus, overall, 87% of the respondents indicated that one or both parents either lost their job or received a cutback in hours (χ^2^ = 53.83, *p* < 0.0001). Among all college dreamers in our sample, 36% reported that they recently got a job or increased their hours at work to provide financial support for their family (χ^2^ = 8.33, *p* = 0.0039), and 24% indicated that they were the breadwinner for their family at some point during the pandemic (χ^2^ = 28.27, *p* < 0.0001).

In terms of mental health, college dreamers who reported that one or both parents recently lost their job due to the COVID-19 pandemic demonstrated greater anxiety (*t*(93) = 2.19, *p* = 0.039) and depression (*t*(92) = 2.79, *p* = 0.0064), and a trend for greater stress (*t*(93) = 1.79, *p* = 0.0768), according to the GAD-7, PHQ-9, and PSS-10 scores. In addition, chi-square analyses revealed that respondents who reported parental job loss were more likely to meet clinical cutoffs for anxiety (χ^2^ = 5.79, *p* = 0.0161) and depression (χ^2^ = 4.29, *p* = 0.0382), suggesting the potential presence of a diagnosable anxiety or depressive disorder. In contrast, there was no evidence of increased mental health issues among dreamers who reported parental cutbacks in hours or for those who recently got a job, increased hours at work, or served as breadwinners for their families during the pandemic.

#### Discussion

A high proportion of the dreamers in our sample reported that their parents faced a change in employment during the COVID-19 pandemic, with 87% indicating at least one parent lost their job or faced a cutback in hours. Moreover, 1 in 3 of the college dreamers in our sample recently got a job or increased hours at work to provide financial support for their family, and nearly 1 in 4 served as the breadwinner for their family at some point during the pandemic. The findings suggest that lapses in household income during the COVID-19 pandemic impelled many undocumented college students to chip in and provide financial support for their families in order to make ends meet. Generally speaking, the parents of dreamers are undocumented immigrants and are thus typically barred from receiving unemployment benefits or other economic relief packages, such as those recently established by the CARES Act. As a result, a drop in income for undocumented parents during the COVID-19 pandemic would have a more dramatic impact on the financial security and, consequently, the mental health of the household. This could, in part, explain the high rate of financial concerns and mental health issues in the present sample of dreamers, especially for those whose parents lost their jobs. On the other hand, it was surprising that college dreamers who worked more and became breadwinners for their families during the pandemic did not suffer a greater mental health impact. However, it should be noted that it is common for dreamers to work while attending college (Teranishi et al., 2015) and to help their families when it comes to finances (Luna & Montoya, 2019). Therefore, to many college dreamers, contributing more to the family during the pandemic may have represented a natural extension of their pre-pandemic behavior, and thus did not signify a major source of added mental health distress.

## General Discussion

The present research was designed to gain a preliminary understanding of the mental health consequences of the COVID-19 pandemic for dreamers currently enrolled in college. While it may be challenging to quickly identify and recruit a large sample of undocumented college students for the purposes of research, the present study employed a fortuitous and readily available population of dreamers currently enrolled in a public university in Delaware. Specifically, the study aimed to (1) identify clinical rates of anxiety, depression, and stress, in addition to general mental health concerns, among the recruited sample of college dreamers during the COVID-19 pandemic, (2) compare these findings with available data from the U.S. general population of college students, and (3) determine whether concerns about education, immigration, and unemployment (among other factors) were associated with greater clinical levels of anxiety, depression, and stress in this sample of dreamers. The findings demonstrate remarkably high clinical levels of anxiety and depression with 47% of dreamers meeting the cutoff for anxiety, 63% meeting the cutoff depression, and 67% meeting the cutoff for anxiety and/or depression, according to the GAD-7 and PHQ-9 scales. While these questionnaires do not serve to diagnose psychological disorders, they demonstrate the high prevalence of anxiety and depressive symptoms in our sample of dreamers. Importantly, those who met the clinical cutoffs would likely meet full diagnosis for an anxiety or depressive disorder upon further evaluation. We also found that 29% of the dreamers in our sample met the cutoff for suicidal ideation (having thoughts of hurting themselves or that they would be “better off dead”), and overall respondents yielded a perceived stress score of 23.26 which was much higher than the typical score expected of 18–29 year old respondents (14.21; Cohen & Williamson, 1988).

While we do not possess pre-pandemic mental health data for the population of dreamers in our study, there is reason to believe that the present findings represent an *increase* in mental health issues as a result of the COVID-19 pandemic. For instance, the proportions of dreamers meeting cutoffs for anxiety and depression in our sample were significantly higher than those observed in prior, pre-pandemic research on undocumented college students (Teranishi et al., 2015; O’Neal et al., 2016; Campos, 2020). In addition, more than half the dreamers in our sample said that the pandemic had a serious negative impact on their mental health, and this group of respondents yielded higher scores for anxiety, depression, and stress, and were also more likely to meet clinical cutoffs for anxiety, depression, and suicidal ideation. Heightened concerns about finances, academics, and immigration, in addition to parental job loss as a result of the pandemic, were also related to higher scores for anxiety and depression (and, in some cases, stress) among the dreamers in our study. A reasonable interpretation of these findings is that the COVID-19 pandemic augmented concerns about finances, academics, immigration, etc., thereby increasing mental health issues in the present sample of dreamers. However, further research and analyses will be required to confirm this interpretation. In particular, longitudinal data drawn at different timepoints would be beneficial for understanding how changes in pandemic-related concerns might contribute to the high rate of mental health problems presently observed in college dreamers.

The present findings also suggest that the COVID-19 pandemic had a stronger negative influence on the mental health of dreamers, compared to the general population. According to an APA poll taken in March 2020, 1 in 3 Americans reported that the pandemic had seriously impacted their mental health, whereas more than half the dreamers in our study reported a similar mental health impact. The proportions of dreamers in our study who met the cutoff for anxiety, depression, and anxiety or depression were significantly higher than the U.S. general population, according to a CDC survey conducted at a similar time to the present survey. In particular, the proportion of dreamers meeting the cutoff for depression was twice the national rate. While younger people and, in particular, college students may be more prone to mental health issues than the U.S. general population, the rate of anxiety and depression observed for the present sample of dreamers was also significantly higher than figures recently reported for college students overall during the pandemic (Healthy Minds Network and American College Health Association, 2020). Thus, the present findings are consistent with predictions from social scientists that the COVID-19 pandemic would have a disproportionately negative influence on the mental health of undocumented immigrants (Clark et al., 2020; Garcini et al., 2020; Page et al., 2020; Wilson & Stimpson, 2020).

While the present study was not designed to uncover the precise *mechanisms* through which the COVID-19 pandemic impaired mental health of college dreamers, the findings provide some potential factors that may be investigated in future work, including heightened concerns about education, immigration, and finances. Most undocumented families are low-income and do not possess the savings or investments to help the household stay afloat following a sudden loss of employment. Moreover, undocumented families are generally barred from receiving government assistance, including unemployment benefits and economic relief plans recently established during the COVID-19 pandemic via the CARES act. For undocumented heads of household who have continued to work through the pandemic, these individuals may be more likely to work essential jobs that put them at greater risk for COVID-19 infection (Page et al., 2020). Furthermore, immigrants in general are more likely to have pre-existing health conditions that potentially make COVID-19 deadlier (Commodore-Mensah et al., 2018). The vast majority of respondents in our sample reported that at least one immediate family did not currently have health insurance, making them less likely to seek or receive medical treatment (Hacker et al., 2015). Economic and health-related challenges faced by undocumented parents have the potential to trickle down to children and harm their mental health (Schaller & Zerpa, 2019; Gassman-Pines, Ananat, & Fitz-Henley, 2020). In our study, the COVID-19 pandemic has not only made undocumented college students worry about their family’s health and finances, but it has also increased their fears of deportation, detainment, and DACA termination. The pandemic has also reportedly hurt their academic performance and made them uncertain about whether they will still be able to achieve their academic and professional goals. Taken together, the pandemic has burdened college dreamers with a heavy load of concerns that have the potential to impair mental health. Further research will be required to determine the extent to which each of these factors is responsible for the high rate of mental health problems facing college dreamers.

The present study represents an important preliminary step in understanding the mental health impact of the COVID-19 pandemic for undocumented college students, while also raising new questions that may be pursued in future work. For instance, a major limitation of the present study was that the sample of college dreamers were fairly homogenous in terms of geography (all respondents attended the same university, and most had permanent residence in North Carolina, Georgia, or South Carolina). Therefore, it will be important for future research to confirm that the high rates of mental health issues observed in the present study would be reflected in a national survey of undocumented college students, while also exploring potential differences between students with DACA vs. those without DACA. Indeed, previous research has indicated that DACA may influence the mental health of undocumented college students (Teranishi et al., 2015). It would also be beneficial for future research to examine whether the pandemic has increased other mental health issues common among undocumented immigrants, including post-traumatic stress disorder, panic disorder, and substance abuse (Garcini, Peña, Galvan, Fagundes, Malcarne, & Klonoff, 2017). As the COVID-19 pandemic continues with no end in sight, it gives researchers the opportunity to explore these and related questions with the overarching aim to learn more about the influence of pandemics and economic crises on the mental health of undocumented college students. It is hoped that this line of investigation will encourage the creation and implementation of better government policies that consider the mental health of this marginalized group during the present COVID-19 pandemic, as well as other health-related and economic crises that are bound to crop up in the future.

## Data Availability

N/A

## Acknowledgments

The authors acknowledge the assistance of Kevin Noriega in serving as the liaison between ourselves and undocumented college students in Delaware.

